# Imaging the effects of tumour necrosis factor inhibition on pain processing in severe rheumatoid arthritis

**DOI:** 10.1101/2023.02.27.23286245

**Authors:** Karolina Wartolowska, Mark Jenkinson, Jesper Andersson, B. Paul Wordsworth, Irene Tracey

## Abstract

**Background:** Monoclonal antibodies against tumour necrosis factor (TNF) markedly reduce inflammation and disease activity in rheumatoid arthritis; however, the mechanisms through which they affect pain are not fully understood.

**Aims:** The aim of this study was to investigate how monoclonal antibodies against TNF alter pain processing and to determine whether neuroimaging can be used as a marker of treatment efficacy and a predictor of treatment response.

**Methods:** Functional magnetic resonance imaging was used to study the neural correlates of clinically-relevant pain evoked by pressing the most painful joint of the right hand and experimental pain evoked by a thermal stimulus applied to the right forearm. A flashing checkerboard was used as a control stimulus. Patients with severe rheumatoid arthritis, qualifying for the anti-TNF treatment, were scanned before the beginning of the therapy and then approximately one and six months after the first injection.

**Results:** TNF inhibition was associated with a marked reduction in pain ratings, inflammation, disease activity as well as depression and catastrophising scores. Effective treatment was linked with less pressure-evoked brain activation in the regions involved in the processing of the sensory aspect of pain and in the limbic structures. Baseline pressure-evoked activation in the thalamus predicted future response to treatment. There was no reduction in heat-evoked brain activation; on the contrary, there was an increase in the activation in the precuneus, which is involved in interoception. There were no differences in response to the visual stimulus.

**Conclusions:** TNF inhibition strongly reduces brain activation in response to clinically relevant pressure pain but not experimental heat pain and these changes reflect the decrease of nociceptive input from the periphery due to the reduction of inflammation as well as central changes in pain modulation. Neuroimaging methods have the potential to explain and predict treatment effects in inflammatory pain conditions.

## 1. Introduction

Rheumatoid arthritis (RA) is a chronic inflammatory disease with a relatively well-understood pathology. Joint pain is the most disabling symptom of RA, and for most patients, it is the main symptom they would like to reduce [1].

Pro-inflammatory cytokines, especially tumour necrosis factor (TNF), play a crucial role in the pathogenesis of RA [2] and are important in the generation of pain [3]. TNF inhibition blocks the pro-inflammatory cytokines cascade and results in a significant improvement in general well-being, pain, joint tenderness, and joint swelling within days from the initiation of treatment [2].

Functional magnetic resonance imaging (FMRI) is a non-invasive technique suitable for investigating the mechanisms involved in the modulation of pain in healthy volunteers [4] and chronic pain conditions [5,6]. Earlier neuroimaging studies on pain processing in arthritis reported increased pressure-evoked brain activation [7] and decreased heat-evoked activation [6] in RA patients. However, some studies did not show any differences in response to heat between arthritis patients and healthy controls [8,9].

TNF inhibition is very effective clinically, but its central neurobiological effects are not fully understood. The aim of this study was to investigate the modulatory effect of anti-TNF treatment on pain processing in RA and to examine whether neuroimaging can be used as a marker of treatment effect and a predictor of treatment response. The hypothesis was that TNF inhibition would reduce brain activation in response to mechanical and thermal stimuli due to decreased inflammation and sensitisation.

## 2. Methods

### 2.1 Patients

Patients with active RA considered for initiation of anti-TNF therapy were consecutively recruited from the Nuffield Orthopaedic Centre, Oxford, UK.

Patients were included if they had seropositive, erosive arthritis and fulfilled the American College of Rheumatology criteria for RA [10]. All participants were previously naïve to anti-TNF medication.

Participants were excluded for the following reasons: A.) any neurological, psychiatric or medical condition that could affect the results of the study other than depression, which is a common co-morbidity in chronic pain [11]; B.) any medication acting on the central nervous system other than low dose antidepressants, such as amitriptyline < 25mg per day; and C.) contraindications for MRI.

### 2.2 Ethics

The study was approved by the Oxfordshire Research Ethics Committee and conducted in accordance with the principles of the Declaration of Helsinki. Written informed consent was obtained from all the participants at the beginning of the first visit.

### 2.3 Treatment

Patients received anti-TNF treatment either alone or in combination with non-biological disease-modifying anti-rheumatic drugs. At the time of the study, there were three TNF inhibitors approved for clinical use in the UK: infliximab (Remicade), etanercept (Enbrel), and adalimumab (Humira). All three have comparable efficacy, despite differences in their structure and their action at the molecular level [12]. Patients were asked not to take non-steroidal anti-inflammatory drugs for 24 hours before each scan.

### 2.4 Study design

This was an observational study of patients with active rheumatoid arthritis treated with anti-TNF drugs. A placebo control was considered unethical due to the severity of the disease and the long duration of the follow-up. The eligibility for the treatment and subsequent response to therapy was assessed by rheumatologists from the Nuffield Orthopaedic Centre. At three months, the rheumatologists classified the patients as responders or non-responders depending on the reduction in the Disease Activity Score in 28 joints with inflammation measured using the erythrocyte sedimentation rate (DAS28-ESR) [17] and low current disease activity [13]. The researchers responsible for the imaging part of this study were not involved in patients’ care and the treating physicians were blinded to the imaging results.

Patients were scanned on three occasions: at the baseline visit before the anti-TNF treatment, at the short-term follow-up visit between two and four weeks after the first injection, and at the long-term follow-up visit between six and ten months after starting the treatment. The time windows for the follow-up visits were chosen based on the results of clinical studies, which have demonstrated a significant reduction in inflammation after two weeks [14] and the maximum effect of treatment usually by six months [15].

At the baseline visit, all participants underwent a comprehensive medical assessment, including co-morbidities and medication. At all three visits, patients were asked to rate their average daily pain intensity (DPI) on an 11-point verbal Numerical Rating Scale (NRS) where 0 represented “no pain” and 10 “the worst pain imaginable” [16]. Disease activity was assessed using the Disease Activity Score in 28 joints (DAS28-ESR) [17], which includes tender joint count, swollen joint count, the erythrocyte sedimentation rate (ESR), and the patient’s subjective rating of their general health. The blood samples for ESR were taken at the end of each visit to avoid the effect of venepuncture pain on the scanning session. The duration of early morning stiffness of the joints (EMS) [18] was also recorded. All participants were asked to complete the Beck Depression Inventory (BDI) [19], and the Pain Catastrophising Scale (PCS) [20] questionnaires, as both depression and catastrophising may influence the pain experience [16].

### 2.5 Stimulation

Mechanical pressure of the most tender joint of the patient’s right hand was used to evoke a clinically relevant pressure pain. This joint was compressed with a purpose-built, MRI-compatible device, which consisted of a 1 cm^2^ rubber probe attached to a spring and a piston. The intensity of the stimulus was identified by using the method of limits [21] so that it reliably evoked moderate pain of 5–6 on the 11-point NRS. This stimulus intensity was the same for all the visits.

Heat-evoked pain was used as a control condition to examine changes in pain processing that were independent of the peripheral disease process. Heat stimulation was delivered using an in-house built, MRI-compatible thermal resistor with a contact area 1.5 × 2 cm and a fast ramp time (from 30 to 60 degrees Celsius in 0.8 s). The thermode was attached to the volar surface of a patient’s right forearm. The pressure and temperature required to evoke moderate pain 5-6 on the NRS were established outside the scanner at the baseline visit using the method of limits and were stimulus-locked for all the visits.

During the scanning session, the pressure and heat stimuli were repeated 10 times (duration 2 s, inter-stimulus interval jittered between 50 and 70 s to avoid the effect of anticipation and habituation). After each scan, subjects were asked to rate the average intensity of pain evoked by the noxious stimulation on the 11-point NRS. After the functional scans with noxious stimulation, the structural scan was acquired for anatomical reference. The imaging session ended with a visual stimulation used as a simple sensory paradigm to assess the non-specific effects of medication on brain activation. The visual stimulus was 10 blocks of black and white checkerboards flickering for 15 s at a frequency of 8 Hz alternated with blocks of rest for 15s, generated using Presentation software v.11.0.

### 2.6 MRI data acquisition

All imaging was performed at the Oxford Centre for Clinical Magnetic Resonance Imaging Research (OCMR) at the John Radcliffe Hospital, Oxford, UK. Data were acquired using a 3T Tim Trio Siemens MR scanner with a single channel head coil. A structural scan was acquired using an MP-RAGE sequence with the following parameters: TR = 2,040 ms, TE = 5.56 ms, TI = 1,100 ms, flip angle 8 degrees, field of view 192×160mm, voxel size 1×1×1 mm.

Functional data were acquired using a standard whole-brain gradient echo EPI sequence with the following parameters: TR = 3 s, TE = 30 ms, flip angle 90degrees, 36 axial slices covering the whole brain, field of view 192 ×256 mm, matrix size 64 ×64, voxel size 3 ×3mm in plane and 3.3 mm slice thickness.

### 2.7 Analysis of clinical, psychological, and psychophysical data

Treatment-related changes between the baseline and the short- and long-term follow-up visits were analysed using the Wilcoxon Signed Ranks test (2-tailed). The Mann-Whitney U test (2-tailed) was used for comparison between patients and controls. Non-parametric tests were chosen because pain ratings were ordinal, and several other measures were non-normally distributed.

### 2.8 Analysis of imaging data

The FMRI data were analysed using FEAT v.5.98, a software tool for model-based FMRI data analysis. FEAT is part of the image analysis package FSL v.4.1.7 (www.fmrib.ox.ac.uk/fsl)[22].

#### 2.8.1 Single subject analysis

The non-brain structures were removed using BET [23]. Then, the data were spatially smoothed with a Gaussian kernel with FWHM of 5 mm and normalised to the same mean intensity by a single scaling factor. A high-pass filter cut-off of 100 s was used to remove low-frequency artefacts. Motion correction was performed with MCFLIRT [24], which aligns all data applying the rigid body transformation with 6 degrees of freedom (DOF). Furthermore, motion artefacts were identified using MELODIC [25] and removed. For each stimulus type, an expected response model was created by convolving the stimulus function with a standard Hemodynamic Response Function and then entered into the General Linear Model (GLM) and fitted to the data. This yielded a set of parameters quantifying the response to each stimulus. Z (Gaussianised T/F) statistic images were thresholded using clusters determined by Z>3.1 and a corrected cluster significance threshold of p=0.05.

#### 2.8.2 Registration

The functional scan was registered to a structural image of each patient using a linear registration with 6 degrees of freedom with FLIRT [24]. The structural image was registered to a standard structural template (MNI152) first using a linear registration with 12 degrees of freedom and then using a non-linear registration method, FNIRT [27]. The functional data were co-registered between the visits and registered non-linearly to the standard space using one common transform to improve the registration between the sessions. This resulted in a better data realignment of data as the fit was similar for all visits.

#### 2.8.3 Higher-level analysis

The higher-level analysis was performed using FLAME2 with automatic outlier detection [28,29].

A tripled paired t-test was performed to assess the effects of treatment on brain activation between the baseline, short-term, and long-term follow-up visit for each type of activation. To investigate the neural correlates of inflammation and pain ratings, the tripled paired t-test was also performed with demeaned values of disease activity score (DAS28-ESR) and pain ratings entered as covariates of interest.

To investigate the predictive value of pain-evoked brain activation, pressure-related baseline scans of responders were compared to the scans of non-responders using an unpaired t-test, with age and sex as covariates of no interest.

Clusters were considered significant at Z threshold above 3.1 and p threshold below 0.05.

## 3. Results

### 3.1 Patients’ characteristics

Twenty-three patients (18 female; median age 63 years, interquartile range - IQR 22; median disease duration 17 years, IQR 38.5) completed the baseline and the short-term visit (Table 1).

**Table 1:**
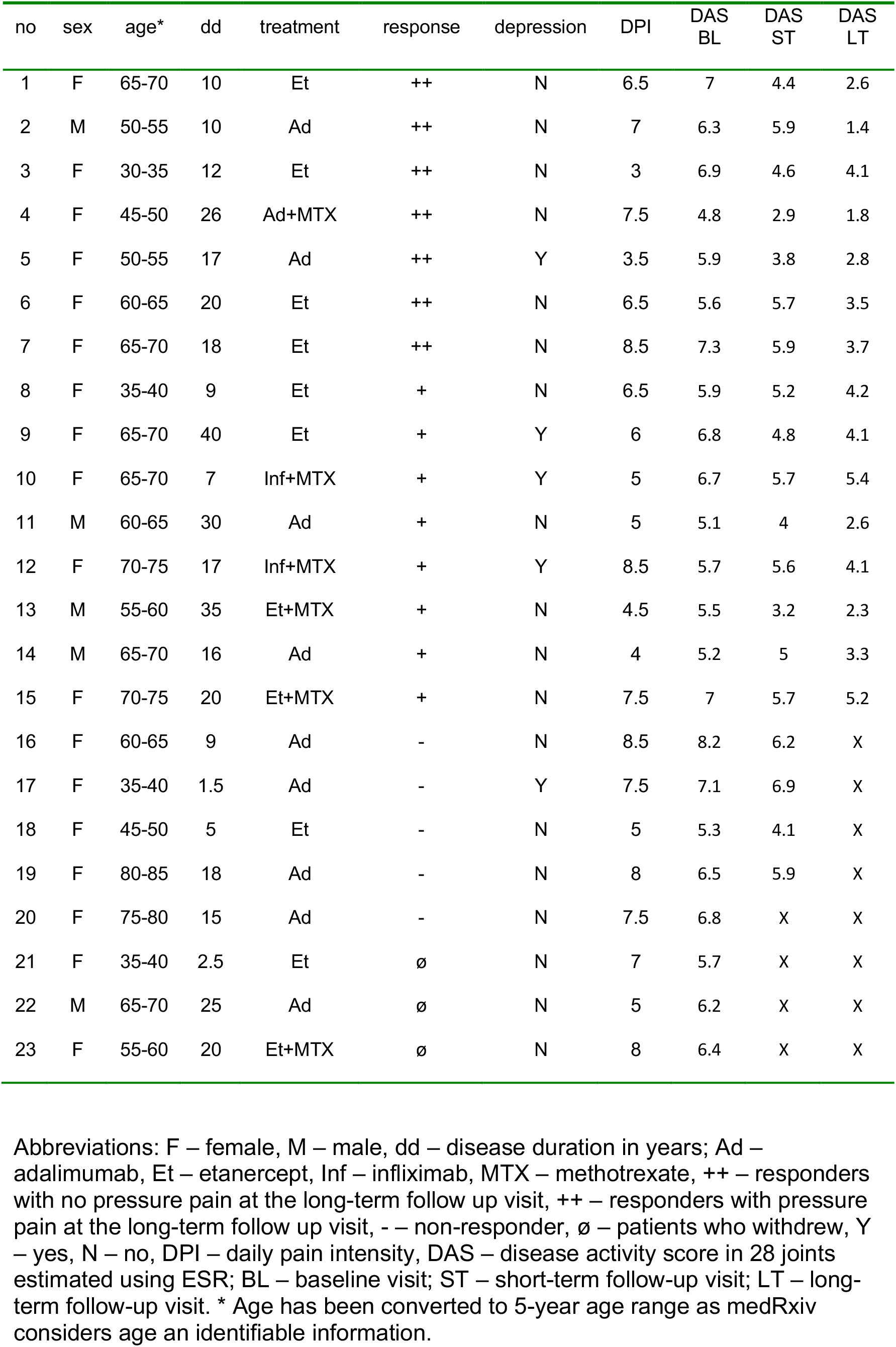
Patients’ characteristics

| no | sex | age* | dd | treatment | response | depression | DPI | DAS<br>BL | DAS<br>ST | DAS<br>LT |
| --- | --- | --- | --- | --- | --- | --- | --- | --- | --- | --- |
| 1 | F | 65-70 | 10 | Et | ++ | N | 6.5 | 7 | 4.4 | 2.6 |
| 2 | M | 50-55 | 10 | Ad | ++ | N | 7 | 6.3 | 5.9 | 1.4 |
| 3 | F | 30-35 | 12 | Et | ++ | N | 3 | 6.9 | 4.6 | 4.1 |
| 4 | F | 45-50 | 26 | Ad+MTX | ++ | N | 7.5 | 4.8 | 2.9 | 1.8 |
| 5 | F | 50-55 | 17 | Ad | ++ | Y | 3.5 | 5.9 | 3.8 | 2.8 |
| 6 | F | 60-65 | 20 | Et | ++ | N | 6.5 | 5.6 | 5.7 | 3.5 |
| 7 | F | 65-70 | 18 | Et | ++ | N | 8.5 | 7.3 | 5.9 | 3.7 |
| 8 | F | 35-40 | 9 | Et | + | N | 6.5 | 5.9 | 5.2 | 4.2 |
| 9 | F | 65-70 | 40 | Et | + | Y | 6 | 6.8 | 4.8 | 4.1 |
| 10 | F | 65-70 | 7 | Inf+MTX | + | Y | 5 | 6.7 | 5.7 | 5.4 |
| 11 | M | 60-65 | 30 | Ad | + | N | 5 | 5.1 | 4 | 2.6 |
| 12 | F | 70-75 | 17 | Inf+MTX | + | Y | 8.5 | 5.7 | 5.6 | 4.1 |
| 13 | M | 55-60 | 35 | Et+MTX | + | N | 4.5 | 5.5 | 3.2 | 2.3 |
| 14 | M | 65-70 | 16 | Ad | + | N | 4 | 5.2 | 5 | 3.3 |
| 15 | F | 70-75 | 20 | Et+MTX | + | N | 7.5 | 7 | 5.7 | 5.2 |
| 16 | F | 60-65 | 9 | Ad | - | N | 8.5 | 8.2 | 6.2 | X |
| 17 | F | 35-40 | 1.5 | Ad | - | Y | 7.5 | 7.1 | 6.9 | X |
| 18 | F | 45-50 | 5 | Et | - | N | 5 | 5.3 | 4.1 | X |
| 19 | F | 80-85 | 18 | Ad | - | N | 8 | 6.5 | 5.9 | X |
| 20 | F | 75-80 | 15 | Ad | - | N | 7.5 | 6.8 | X | X |
| 21 | F | 35-40 | 2.5 | Et | ø | N | 7 | 5.7 | X | X |
| 22 | M | 65-70 | 25 | Ad | ø | N | 5 | 6.2 | X | X |
| 23 | F | 55-60 | 20 | Et+MTX | ø | N | 8 | 6.4 | X | X |

Abbreviations: F – female, M – male, dd – disease duration in years; Ad – adalimumab, Et – etanercept, Inf – infliximab, MTX – methotrexate, ++ – responders with no pressure pain at the long-term follow up visit, ++ – responders with pressure pain at the long-term follow up visit, - – non-responder, ø – patients who withdrew, Y – yes, N – no, DPI – daily pain intensity, DAS – disease activity score in 28 joints estimated using ESR; BL – baseline visit; ST – short-term follow-up visit; LT – long-term follow-up visit. \* Age has been converted to 5-year age range as medRxiv considers age an identifiable information.

All patients were on stable doses of the non-biological disease-modifying anti-rheumatic drugs during the time of the study, except one patient, whose dose of leflunomide was reduced, and one who had their leflunomide stopped. One patient was diagnosed with depression and was treated with amitriptyline at stable doses. All patients scored over 26 on the Mini Mental State Examination and did not show any signs of cognitive impairment.

Between the short-term and the long-term visit, five patients did not respond to the prescribed anti-TNF therapy and had the anti-TNF medication stopped or changed by their treating physician. Patients who did not respond had a higher tender joint count and longer early morning joint stiffness at baseline (the Mann–Whitney test Z=-2.4, p=0.015 and Z=-2.1 p=0.04, respectively).

Three patients withdrew from the study for non-clinical reasons.

The remaining 15 patients (11 female; median age 64 years, IQR 18; median disease duration 17 years, IQR 16) responded to the original anti-TNF treatment and attended all three visits. In this group, the median time between the baseline and the short-term visit was 18 days (IQR 7), and between the baseline and the long-term visit was six months (IQR 4).

### 3.2 Treatment effects on clinical and psychological measures

In the 15 responders all outcome measures decreased significantly with anti-TNF treatment (Table 2). Between the baseline and the short-term visit, there was a reduction in all the measures, except for the catastrophising. Between the short-term and the long-term visit, there was a further marked decrease in all the measures, except for the pain ratings for pressure and heat, swollen joint count, and the duration early morning stiffness, which did not reach significance.

**Table 2:** Clinical, psychological, and psychophysical measures at each visit

| Variable | BL<br>*Median (IQR)* | ST<br>*Median (IQR)* | LT<br>*Median (IQR)* | BL vs. ST<br>*p value* | ST vs. LT<br>*p value* | BL vs. LT<br>*p value* |
| --- | --- | --- | --- | --- | --- | --- |
| DAS28-ESR | 5.9 (1.4) | 5.0 (1.7) | 3.5 (1.5) | 0.001 | 0.001 | 0.001 |
| TJC | 11.0 (7.0) | 10.0 (8.0) | 4.0 (8.0) | 0.005 | 0.002 | 0.001 |
| SJC | 11.0 (7.0) | 8.0 (5.0) | 3.0 (5.0) | 0.004 | 0.07 | 0.001 |
| ESR, mm/h | 34 (29) | 23 (23) | 13 (11) | 0.008 | 0.002 | 0.001 |
| DPI | 6.5 (3.0) | 4.0 (3.0) | 2.5 (3.5) | 0.004 | 0.002 | 0.001 |
| GH | 70 (27) | 35 (40) | 20 (20) | 0.001 | 0.01 | 0.001 |
| EMS, min | 54.7 (45.0) | 10.0 (60.0) | 0.0 (20.0) | 0.005 | 0.09 | 0.001 |
| PRp | 5.0 (1.1) | 3.0 (3.2) | 2.0 (3.1) | 0.002 | 0.07 | 0.001 |
| PRh | 6.0 (0.5) | 5.5 (2.0) | 5.5 (2.0) | 0.006 | 0.4 | 0.1 |
| BDI | 8.0 (6.0) | 6.0 (9.0) | 4.0 (5.0) | 0.02 | 0.03 | 0.001 |
| PCS | 10.0 (18.0) | 11.0 (9.0) | 3.0 (5.0) | 0.06 | 0.003 | 0.005 |

Absolute values (median and interquartile range) at each visit and significance of changes between visits (*p* values of the Wilcoxon Signed Ranks test) for clinical, psychophysical and psychological measures in the 15 responders who completed all three visits. Abbreviations: BL – baseline visit; ST – short-term follow-up visit; LT – long-term follow-up visit; DAS28-ESR – disease activity in 28 joints; TJC – tender joint count; SJC – swollen joint count; ESR – erythrocyte sedimentation rate; DPI – daily pain intensity; GH – the global impact of RA on general health; EMS – duration of early morning joint stiffness; PRp – pain intensity of the pressure stimulus; PRh – pain intensity of the heat stimulus; BDI – Beck Depression Inventory; PCS – Pain Catastrophising Scale.

Between the baseline and the short-term visit, the reduction in disease activity measured with DAS28-ESR correlated positively with measures of perceived improvement, such as the Patients’ Global Impression of Change and a change in general health (Spearman’s rho=0.69 p=0.004 and rho=0.74 p=0.02, respectively). There was also a positive correlation between a reduction in an inflammatory marker, ESR, and a reduction in the depression score (rho=0.72 p=0.002). The decrease in pain ratings for pressure and heat did not correlate with changes in any clinical or psychological measures at the short-term visit; however, there was a correlation between a reduction in DAS28-ESR and in pressure-pain ratings (rho=0.58 p=0.02) between the baseline and the long-term visit.

The pressure stimulus was no longer painful (PR < or = 1) for three patients at the short-term visit and for further four patients at the long-term follow-up visit. The only differences between the eight patients with pain and seven without pain, in the pressure-pain ratings at the long-term visit (the Mann-Whitney test Z=-3.3 p<0.0005) and in the reduction in the pressure-pain ratings and DAS28-ESR between the baseline and the long-term visit (the Mann-Whitney test Z=-3.2 p=0.001 and Z=-2.5 p=0.009, respectively).

### 3.3 Treatment effect on brain activation in 15 responders

#### 3.3.1 Pressure-evoked activation

Between the baseline and the long-term visit, anti-TNF medication was associated with a reduction in pressure-pain evoked activation in the left primary sensorimotor cortex, bilaterally in the hippocampi and the right amygdala (Figure 1 and Supplementary Table 1). This effect was driven by the seven patients who reported no pain on joint stimulation at the long-term visit (Supplementary Figure 1 and Supplementary Table 2).

**Figure 1.**
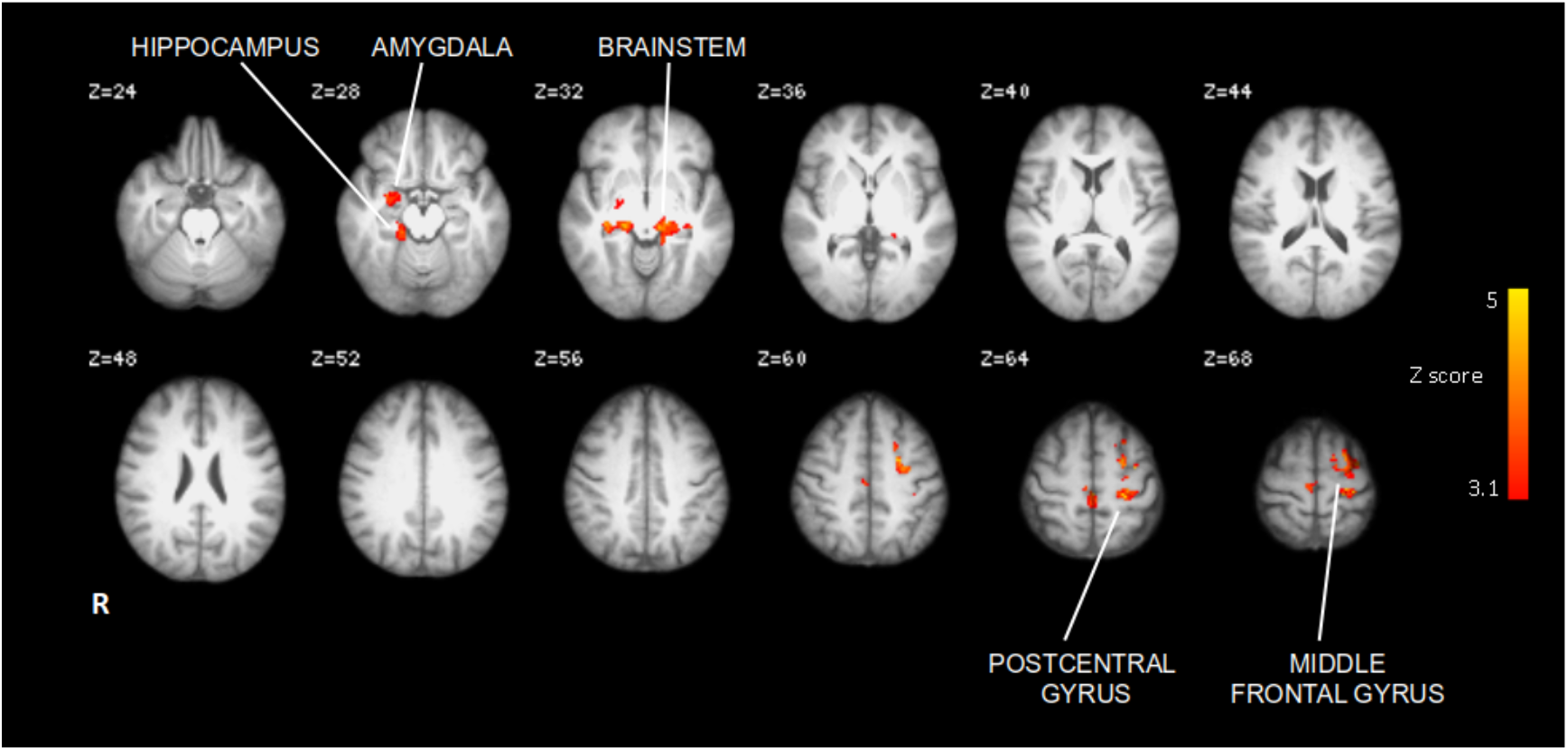
Reduction of activation in response to pressure stimulus between baseline and the long-term visit in all 15 responders. Changes were present in the right amygdala, bilaterally in the hippocampus, and the left primary somatosensory cortex. FLAME2, triple paired t-test Z>3.1 p<0.005.

The number of patients was not sufficient to detect significant changes at the short-term visit or correlations between changes in the pressure-evoked activation and changes in DAS28-ESR or pain ratings.

#### 3.3.2 Heat-evoked activation

There was no reduction in heat-evoked brain activation between baseline and the short-term or the long-term visit; however, there was an increase of activation between the baseline and the long-term visit in the precuneus/posterior cingulate cortex (MNI coordinates of the maximum voxel: 2 -52 42). The number of participants was not sufficient to detect correlations between changes in brain activation and DAS28-ESR or pain rating changes.

#### 3.3.3 Visual task-related activation

There were no differences in the activation in response to the visual task after the treatment, suggesting that anti-TNF did not have a global effect on brain activation.

### 3.4 Predictive value of neuroimaging

At baseline, pressure-evoked activation was stronger in the right thalamus (maximum voxel’s MNI coordinates: 20 -28 -4) in the 15 responders than five non-responders (unpaired 2 group tests with age and sex as covariates of no interest).

## 4. Discussion

### 4.1 Main findings

Anti-TNF treatment was associated with a marked decrease in pain, inflammation, and depression as well as a reduction in joint pain-related brain activation in the sensorimotor and limbic regions. There were also baseline differences in pressure-evoked activation in the thalamus between patients who responded and those who did not responded to the anti-TNF therapy. There was also an increase of heat-evoked brain activation in the precuneus but there were no differences in response to the visual control task.

### 4.2 Treatment effect on clinical, psychological, and psychobehavioural outcomes

The significant reduction in disease activity measured using DAS28-ESR, inflammatory markers and pain ratings, observed within the first month of anti-TNF treatment, is consistent with the results of earlier clinical studies [14].

The effect of anti-TNF treatment on joint-related pain is probably mediated mainly through its anti-inflammatory effect in the periphery (i.e. a decrease in swelling and tenderness of the stimulated joint) but may also have a systemic component (i.e. a reversal of inflammation-related augmentation of pain) [14]. Moreover, some of the effects of anti-TNF treatment may be related to its direct action on nerve fibres in the peripheral or the central nervous system [7,32]. The results of this study suggest these mechanisms may be involved in reduction of mechanical hyperalgesia after anti-TNF treatment. Pressure pain ratings decreased within the first month, with the change between the short-term and the long-term visit not reaching significance. This reflects the pattern of changes in the swollen joint count, which is a measure of inflamed joint tissue and effusion [34], supporting the role of the reduction of inflammation in the periphery. Also in the first month there was a marked decrease in the duration of early morning stiffness, which is driven by the disease-related changes in the central nervous system; therefore, pointing towards the anti-TNF on central mechanisms. Finally, pain ratings decreased early in the treatment but there was only a small further reduction at the long-term visit, despite the fact that the DAS28-ESR and ESR further decreased. Moreover, changes in pain ratings did not correlate with a reduction in inflammation. This would suggest that the some of the effect is not mediated through inflammation but may be directly altering pain processing in the periphery or centrally.

It is important to acknowledge that anti-TNF treatment was associated with reduction in depression and catastrophising, which potentially affected patient-reported outcomes such as pain ratings. Due to the small study size, it was not possible to fully disentangle the complex effects of blocking TNF on joint pain.

Finally, some of the effect on the reported measures might have been caused by a placebo effect rather than the clinical effect of the treatment [31]. For example, the reduction in heat pain ratings at the short-term visit might have been caused by the placebo effect. The potential placebo effect could not have been investigated due to the observational nature of this study.

### 4.3 Treatment effect on pain-related brain activation

Reduction in pressure-pain related activity was present in the primary sensorimotor cortex and in the limbic structures. The primary somatosensory cortex is involved engaged in the representation of touch and pain [33], including both the discriminatory component of a painful stimulus and the perceived stimulus intensity [35]. Moreover, responders demonstrated more extensive pressure-evoked brain activation at baseline in the right thalamus, which is a region involved in processing of clinical pain [36] that has been reported to respond to pressure pain in patients with fibromyalgia [38]. Further investigation of these effects could help to identify the subgroup of patients with rheumatoid arthritis who appear to have a prominent element of fibromyalgia to their symptoms.

The treatment-related effect in the amygdala and hippocampus is particularly interesting as these regions are involved in the processing of the affective dimension of pain [40] and their activation is typically reported in studies on anxiety or fear [41]. We could not investigate the effects of anxiety, as it was not specifically assessed in this study. However, we observed that the changes in the limbic regions became evident only later in the treatment and were present for the pressure-evoked but not for the heat-evoked pain. These changes were mostly driven by patients without pain at the long-term visit for whom the stimulus was not only qualitatively different, i.e., less painful, but also quantitatively different, i.e., not painful and not unpleasant. Therefore, we suggest that the observed effects are specific for the pressure-evoked pain and are probably related to the stimulus not being painful and therefore, no longer threatening for patients, rather than the reduction of anxiety related to repeated scanning or learning effect at the follow-up visit [42]. In addition to that, the limbic regions mediate the effects of proinflammatory cytokines on mood [43]. Correlations between activation in the amygdala and depression have been described in subjects with fibromyalgia [37] but were not apparent in our study. Finally, the significant reduction of pressure-evoked signal in the amygdala may also be related to improvement in a more general sense. The amygdala is involved in processing sensory-discriminative aspects of pain, complex pain behaviour, autonomic responses as well as in descending pain modulation, especially during chronic inflammation [44-47]. Activation in this region reflects changes in arthritic pain and pain behaviour after treatment [8,46].

Interestingly, we did not observe a marked decrease in pain ratings or in brain activation in response to thermal stimuli. This suggests that anti-TNF treatment and a reduction in inflammation do not change the processing of heat pain. The results of neuroimaging studies on heat pain in pain conditions are inconsistent. Increased responses to thermal stimuli have been reported in patients with fibromyalgia but not in patients with lower back pain or arthritis [8,48,38]. In contrast, Jones and Derbyshire [6] reported less extensive heat-evoked brain activation in RA patients and suggested that pain processing is altered by ongoing inflammation. In our study, there was an increase in activation after treatment that was present mainly in the regions typically engaged in processing of attention, self-awareness, and integration of sensory information in relevance to self [49]. The posterior cingulate is also involved in pain processing in chronic pain states; however, it encodes the valence of stimuli rather than nociception [50]. The increase in heat-evoked brain activation may be a result of patients paying more attention to the experimental pain and perceiving it as more self-relevant when the clinical pain decreases after treatment; therefore, reflecting an increase of cognitive resources available to perceive the experimental stimulus. These changes seem to be specific to the thermal pain as this effect was not present for the pressure or visual stimulation.

## 5. Conclusions

This study demonstrated that TNF has different effects on the processing of clinically relevant pressure-evoked pain and experimental heat-evoked pain. The effect of treatment on mechanical pain processing was present in the somatosensory and limbic structures reflecting changes in the pain input from the periphery as well as changes in central pain modulation.

## Data Availability

All data produced in the present study are available upon reasonable request to the authors.

## 6. Acknowledgment and Conflict of Interest Statement

This research project was supported by unrestricted grants from GlaxoSmithKline (KW), the Wellcome Trust (IT), the Biotechnology and Biological Sciences Research Council - David Phillips Fellowship (MJ) and the Medical Research Council of Great Britain and Northern Ireland (FMRIB Centre). There was no conflict of interest. The funding bodies had no role in study design, data collection and analysis, decision to publish, or preparation of the manuscript.

## 8. Appendix

**Supplementary Table 1.**
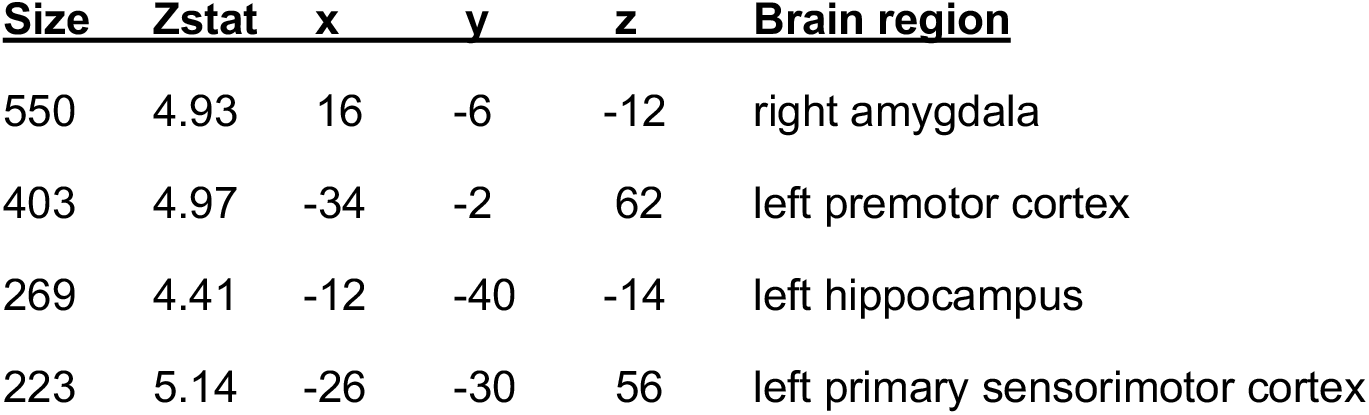
Activation clusters in Figure 1.

**Supplementary Figure 1.**
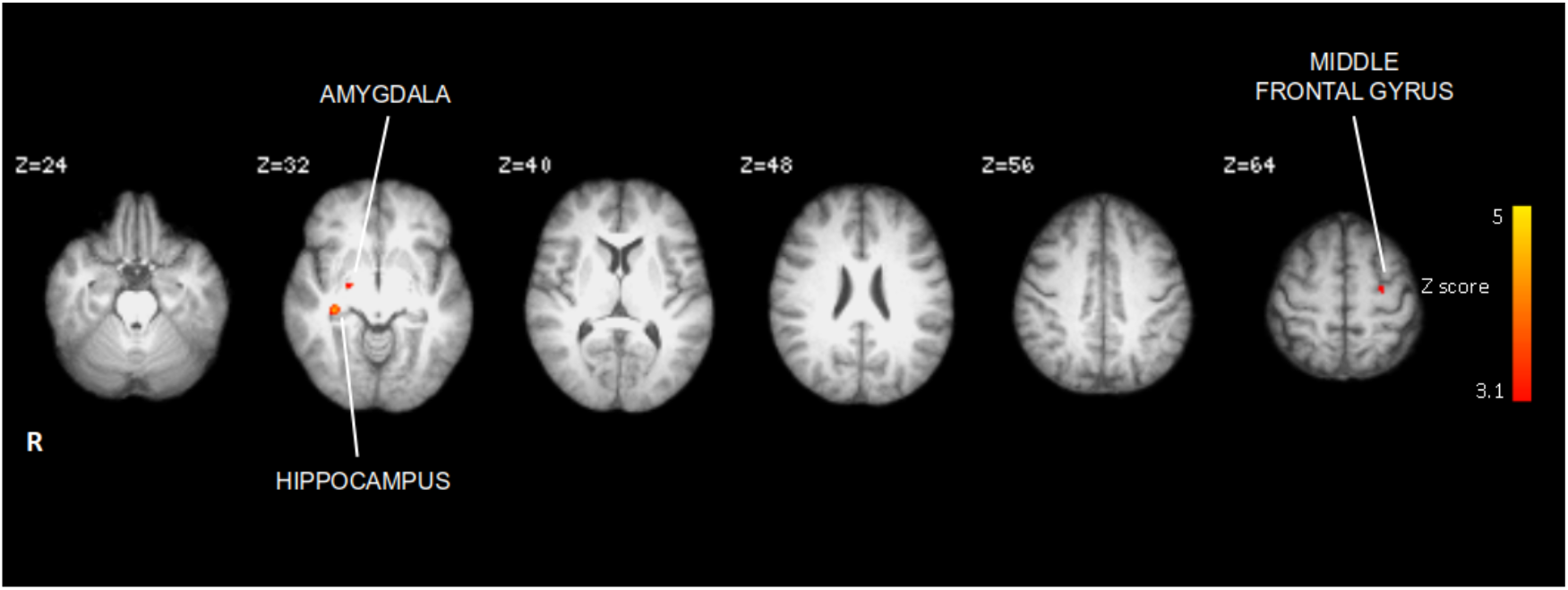
A decrease in activation in response to mechanical stimulation between the baseline and the long-term visit in the seven patients with no pain at the follow-up. FLAME2, paired t-test, Z>3.1 and p<0.05.

**Supplementary Table 2.** Activation clusters in Supplementary Figure 1.

| **Size** | **Zstat** | **x** | **y** | **z** | **Brain region** |
| --- | --- | --- | --- | --- | --- |
| 186 | 4.58 | 24 | -8 | -12 | right amygdala |
| 110 | 4.48 | -26 | -10 | 50 | left primary sensory cortex |

